# Class 1 integrons in clinical and swine industry isolates of *Salmonella* Typhimurium from Colombia, dating 1997 to 2017

**DOI:** 10.1101/2022.02.14.22270819

**Authors:** Nancy Yaneth Flórez-Delgado, Elizabeth Noelia Ubillus, Blanca Perez-Sepulveda, Eyda Lizeth Ospina-Ríos, Ana Karina Carrascal-Camacho, Iliana C Chamorro-Tobar, Lucy Angeline Montaño, Yan Li, Silvana Zapata-Bedoya, Jay C.D. Hinton, José Miguel Villarreal, Magdalena Wiesner

**Affiliations:** Grupo de Microbiología, Instituto Nacional de Salud, Avenida calle 26 No. 51-20 - Zona 6 CAN. Bogotá, D.C., Colombia; Departamento de Química, Facultad de Ciencias, Universidad Nacional de Colombia, Carrera 45 # 26-85, Edif. Uriel Gutiérrez, Bogotá D.C., Colombia; Grupo de Investigación en Ciencias Biológicas y Químicas, Facultad de Ciencias, Universidad Antonio Nariño, Calle 58a # 37 – 94, Bogotá, D.C., Colombia; Doctorado en Ciencias de la Salud, Facultad de Medicina, Universidad Antonio Nariño, Calle 58a # 37 – 94, Bogotá, D.C., Colombia; Institute of Infection, Veterinary and Ecological Sciences, University of Liverpool, Biosciences Building, Liverpool, L69 7ZB, United Kingdom; Laboratorio de Microbiología de Alimentos, Semillero de Inocuidad Alimentaria, Grupo de Biotecnología Ambiental e Industrial, Pontificia Universidad Javeriana, Carrera 7 No. 40 – 62, Bogotá D.C., Colombia; Dirección de Investigación en Salud Pública, Instituto Nacional de Salud, Avenida calle 26 No. 51-20 - Zona 6 CAN. Bogotá, D.C., Colombia.

**Keywords:** *Salmonella* Typhimurium, integrons, drug resistance, mobile genetic element, plasmid, Colombia

## Abstract

**Background:** *Salmonella enterica* subsp. *enterica* serovar Typhimurium (*S.* Typhimurium) has been linked to outbreaks of foodborne gastroenteritis disease, and the emergence of antimicrobial-resistant clones. In Colombia, laboratory surveillance of *Salmonella* spp. between 1997-2018 revealed that *S*. Typhimurium was the most ubiquitous serovar (27.57% of all *Salmonella* isolates), with increasing levels of antimicrobial resistance to several families of antibiotics. *Hypothesis*. Resistant isolates of *S.* Typhimurium recovered from clinical and swine samples carry class 1 integrons that are linked to antimicrobial resistance genes. *Aim*. Identify class 1 integrons, and investigate their association with other mobile genetic elements, and their relationship to the antimicrobial resistance of Colombian *S.* Typhimurium isolates. *Methods.* In this study, 392 clinical isolates of *S.* Typhimurium were analyzed, of which 237 were obtained from blood culture, 155 from non-invasive sources and 50 isolates from swine of which 32 were obtained from a slaughterhouse, 10 from a point of sale and 8 from cutting plant. Class 1 integrons and plasmid incompatibility groups were analyzed by PCR and Whole Genome Sequencing (WGS), and the region surrounding of the integrons identified by WGS. The phylogenetic relationship was established by MLST and SNP analysis. *Results.* Overall, 39.03% (153/392) of the clinical isolates and 22% (11/50) of the swine *S.* Typhimurium isolates carried complete class 1 integrons. Twelve types of gene cassette arrays were identified, including *dfr7*-*aac*-*bla*_OXA-2_ (Int1-Col1) as the most frequent in clinical isolates (75.2% = 115/153). Clinical and swine isolates that carried class 1 integrons were resistant to up to 5 and up to 3 antimicrobial families, respectively. The Int1-Col1 integron was most prevalent in stool isolates and was associated with Tn21. The most common plasmid incompatibility group was IncA/C. *Conclusions.* The widespread presence of the integron (IntI1-Col1) in Colombia since 1997 was striking. We speculate that the relationship between integrons, source and mobile elements favors the dispersion of antibiotic resistance determinants in Colombian *S*. Typhimurium isolates.

## Introduction

*Salmonella enterica* is a widely studied pathogen with 2,659 serovars (Issenhuth-Jeanjean *et al*., 2014). Globally in 2017, Non-typhoidal serovars have caused 50,800 deaths, and non-typhoidal invasive *Salmonella* caused 77,500 deaths (GBD 2017 Non-Typhoidal Salmonella Invasive Disease Collaborators, 2019), they are considered one of the main biological hazards that affect food safety (Organización de las Naciones Unidas para la Alimentación y la Agricultura & Organización Mundial de la Salud, 2016). Salmonellosis usually causes gastroenteritis, which can be lethal in children, the elderly, and immunosuppressed patients (Farthing *et al*., 2013). One of the main serovars associated with this disease is *Salmonella enterica* subsp. *enterica* serovar Typhimurium (*S.* Typhimurium) (Hendriksen *et al*., 2011), a serotype that can infect animal and human hosts (Hoelzer *et al*., 2011; Simpson *et al*., 2018). *S.* Typhimurium has been the second most commonly reported serotype in humans samples and was the most reported serovar isolated from cattle, pigs, pork and turkey meat in 2016 in the European Union (European Food Safety Authority & European Centre for Disease Prevention and Control, 2017). Similarly, the laboratory-based National *Salmonella* Surveillance System from US states reported *S.* Typhimurium within the top-five most frequent serovar during 1996-2011 (Boore *et al*., 2015).

In recent decades, the increase and dissemination of antimicrobial resistant *S.* Typhimurium strains has been reported in humans, animals and food (Kingsley *et al*., 2009; Tran-Dien *et al*., 2018). Antimicrobial resistance is a worldwide public health problem (Hu *et al*., 2020; World Health Organization, 2015). The acquisition of pre-existing resistance determinants is achieved through mobile genetic elements such as insertion sequences (IS), transposons (Tn) and gene cassettes/integrons. Integrons can mobilize genes between defined sites using site-specific recombination, forming a cassette array that may confer multi-resistance, which allows the bacteria an adaptation capability to environmental pressures. Integron mobility has been associated with transposable elements and conjugative plasmids (Escudero *et al*., 2015; Hall, 2012; Partridge *et al*., 2018). Mobile genetic elements that promote intra and inter-species intercellular exchanges are plasmids and integrative conjugative elements that facilitate horizontal gene transfer, promoting the acquisition and dispersal of antibiotic resistance genes (Partridge *et al*., 2018).

In Colombia, *S*. Typhimurium has also been one of the most predominant serovars isolated from human samples since 1997 (Instituto Nacional de Salud, 2019; Rodríguez *et al*., 2017). The presence of class 1 integrons was identified in few multidrug resistant (MDR) *S.* Anatum and *S.* Typhimurium isolates from food (O’Mahony *et al*., 2006). However, confirmation, frequency, and distribution of Class 1 integrons in clinical and swine isolates of *S.* Typhimurium and their possible association with increased antimicrobial resistance is unknown.

This work aimed to identify the presence of integrons, and other mobile genetic elements such as IS and plasmids, which favor the acquisition and dispersion of antimicrobial resistance in Colombian isolates of *S.* Typhimurium of human and animal origin between 1997 and 2017.

## Methods

### *S.* Typhimurium clinical isolates

*S.* Typhimurium isolates recovered from clinical samples in 21 departments and the Capital District were characterized for phenotypic and molecular methods under the acute diarrheal disease surveillance program (Instituto Nacional de Salud, 2017). *Salmonella* spp. isolates were serotype by Kauffman-White-Le Minor scheme (Grimont & Weill, 2007) and the antimicrobial susceptibility profile was determined by the disc diffusion method (Kirby-Bauer) following the interpretation criteria of the Clinical & Laboratory Standards Institute (CLSI) M100-S27 (Clinical and Laboratory Standards Institute (CLSI), 2017) for tetracycline (TET), chloramphenicol (CHL), nalidixic acid (NAL), amoxicillin/clavulanic acid (AMC), aztreonam (AZT), amikacin (AMK) and streptomycin (STR). The minimum inhibitory concentration (MIC) was determined in the AutoSCAN-4 (Siemens, Germany) with the panel NC50 ampicillin (AMP), sulfamethoxazole/SXT (SXT), cefotaxime (CTX) and ceftazidime (CAZ). Isolates with resistance to 3 or more antibiotic families were defined as multidrug-resistant (MDR) (Magiorakos *et al*., 2012; Rodríguez *et al*., 2017).

Of this surveillance 392 clinical isolates of *S.* Typhimurium were selected for this study. Among them, 239 (60.97%) strains were isolated from systemic sources: 237 (60.46 %) of blood culture and 2 (0.51 %) of cerebrospinal fluid. One hundred and forty-eight (37.76 %) strains were isolated from non-systemic sources: 137 (34.95 %) of stool; 11 (2.81 %) of other samples. Four (1.02 %) strains were isolated from food sources and one (0.26%) strain do not register source. The isolates were resistant from 1 to 5 antimicrobial families with a total of 51 antimicrobial-resistant profiles, most of them were multidrug-resistant profiles (see Table S1 and the data published by Li et. al., 2019 (S1 Table-metadata of the samples (Li *et al*., 2019)).

### *S.* Typhimurium isolated from animals from a swine industry

The 409 isolates of *Salmonella* spp. found in slaughterhouse, cutting plant and point sale was collected between 2013 to 2015. *Salmonella* was identified by method ISO 6579 (2002) and MDS (Molecular Detection System -3M) (Bird *et al*., 2016). Antimicrobial resistance was determined using Panel B1016-180 (Beckman Coulter, Negative Combo 72, NC72), based on specifications by the Clinical & Laboratory Standards Institute (CLSI) M100-S27 (Clinical and Laboratory Standards Institute (CLSI), 2017).

73 isolates presenting multiple resistances to two or more first or second choice antimicrobials for human salmonellosis treatments were serotyped employing Kauffman-White-Le Minor scheme (Grimont & Weill, 2007). A total of 50 strains that were identified as *S*. Typhimurium were included. Of these isolated, 32 were obtained in the slaughterhouse (19 carcass, 4 knife, 4 saw and 5 other samples), 8 in cutting plants (4 carcass, 2 utensils, and 2 other samples) and 10 in the point of sale (4 meat pulp and 6 other samples) (Table S2).

### DNA extraction of clinical and animal isolates

All bacterial isolates were grown in 200 µl of Brain Heart Infusion (BHI) agar at 37°C overnight (12 to 18 hours). Total genomic DNA was extracted using a commercial kit QIAamp DNA mini-Kit (QIAGEN - Sample & Assay Technologies) for clinical samples, and a commercial kit Wizard Genomic DNA Purification Kit (Promega) for animal/swine samples, following the manufacturer’s protocols. The DNA concentration and quality was measured using a NanoDrop^TM^ 2000c spectrophotometer (Thermo Scientific™), following the manufacturer’s recommendations, and visualized by agarose-gel electrophoresis.

### Determination of class 1 integrons

Search of class 1 integron in Typhimurium isolates recovered in the 392 clinical and 50 swine samples was developed in two phases. First phase included 166 clinical and 50 swine industry isolates, which were tested by conventional PCR and sanger sequencing of the amplified fragments. Second phase included 226 clinical isolates analyzed by whole genome sequencing (WGS). <u>First phase: Determination of class 1 integrons by PCR amplification and sanger sequencing</u>. The genomic DNA of 166 clinical and 50 isolates from swine industry samples, respectively, were used as a template in the PCR, to determine the presence of class 1 integrons. Amplification of *intI*1 (class 1 integrase gene), *qacE*Δ1 (quaternary ammonium compound gene), *sul*1 (dihydropteroate synthase gene) genes and the variable regions (CS’) of class 1 integrons were performed with the primers described in Table 1. It was considered as a complete class 1 integron (complete IntI1), that contains the structural genes *intI1*, *qacEΔ1* and *sul1*. The sequence of the CS’ region was identified by nested PCR. The PCR reaction was prepared with final concentrations of 1X Buffer, 1.5 mM MgCl2, primers that correspond to the *intI1*, CS’ region, *qacEΔ1* and *sul1* genes have a final concentration/tube from 0.24, 0.5, 0.24 and 0.2 µM, respectively; 1U Platinum Taq DNA Polymerase enzyme (Invitrogen), in 25 µl final reaction volume. Product sizes were verified by 1% agarose gel electrophoresis in 1X TAE buffer, at 100 V for 60 min. The PCR products obtained from the CS’ region of the class 1 integron were purified with the DNA Clean & ConcentratorTM-5 Kit (Zymo Research) and were sequenced on ABI 3730xl (Applied Biosystems) sequencers (Macrogen, Korea). The putative resistance genes in the variable region were identified using BLAST (McGinnis & Madden, 2004), using a cut off of 97% or more for determine similarity to the gene reported in the BLASTn database (Johnson *et al*., 2008). <u>Second phase: Determination of class 1 integrons by WGS</u>. Genomic DNA extraction, library preparation and WGS for 225 clinical *S.* Typhimurium isolates were performed as part of the 10KSG consortium and were described previously (Perez-Sepulveda *et al*., 2020). For 9 clinical isolates the WGS was made with Miseq Illumina platform. *De Novo* assembly were performed with CLC Genomics Workbench (Qiagen) version 8.0.1. “Rapid Annotation using Subsystem Technology, RAST” version 2.0 and SEED Viewer (http://rast.nmpdr.org/seedviewer.cgi) were used for annotation of genomes and with the pipeline available from https://github.com/apredeus/10k_genomes (Perez-Sepulveda *et al*., 2020).

**Table 1.**
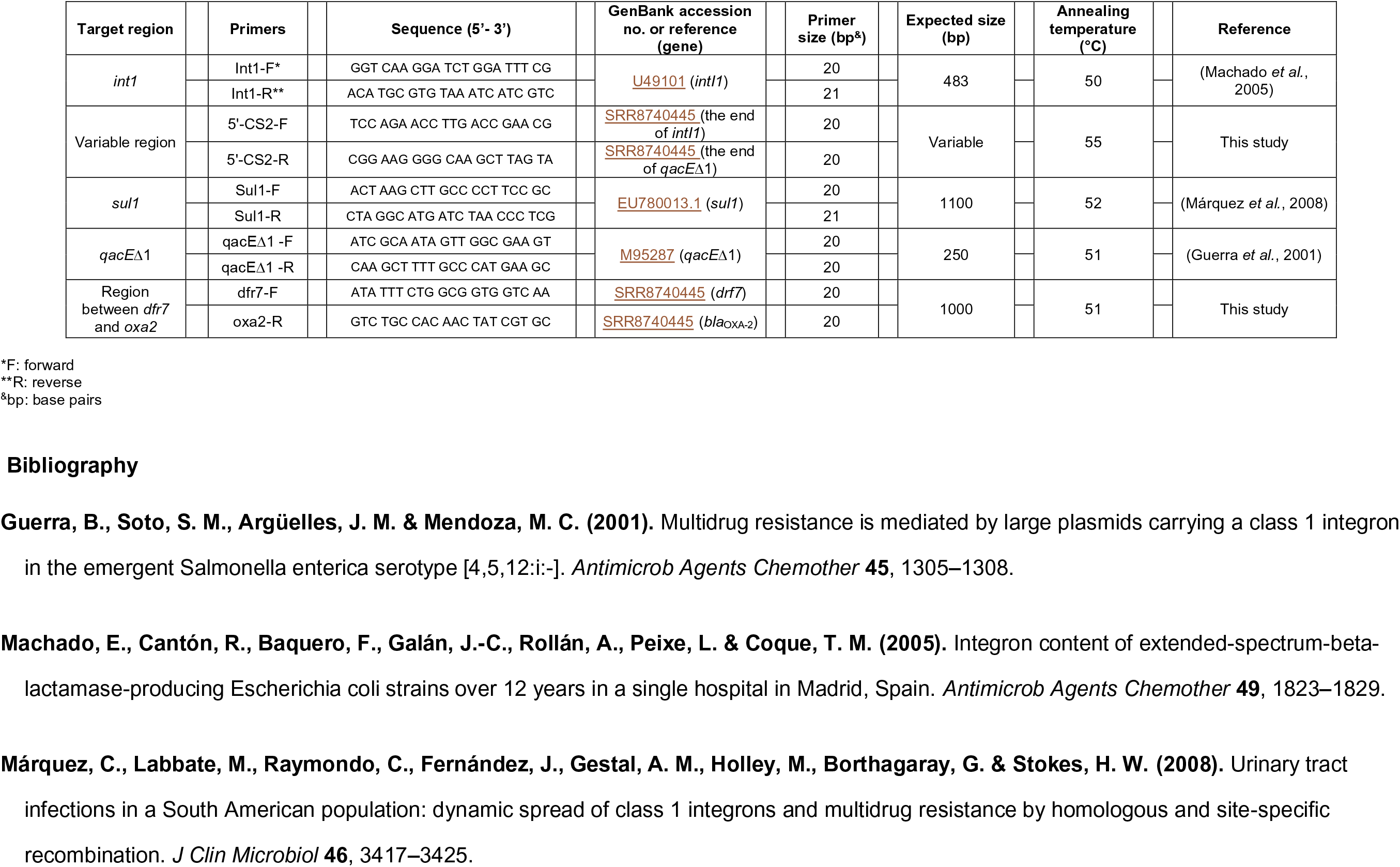
PCR primers for class 1 and 2 integrons.

### Sequence analysis and class 1 integrons search

Annotation of assembled genomes was carried out with PATRIC 3.5.43 (Wattam *et al*., 2017), using the keywords: integron integrase, qacE, delta 1, dihydropteroate synthase type-2 (sul1) and, variable region resistance genes that were identified by sequencing of PCR products. The genome annotation in PATRIC was performed comparing external databases: CARD (Comprehensive Antibiotic Resistance Database) (Jia *et al*., 2017)), DrugBank (a database with molecular information, mechanisms, interactions and drug targets) (Wishart *et al*., 2018), NDARO (National Database of Antibiotic Resistant Organism) and PATRIC (Wattam *et al*., 2017). The results obtained were confirmed in the BLAST (McGinnis & Madden, 2004) and INTEGRALL (Moura *et al*., 2009) databases.

### Surrounding regions of the class 1 integrons

We used the genome sequence obtained in *S*. Typhimurium clinical isolates positive for class 1 integrons, to reconstruct the surrounding regions and identify other mobile genetic elements flanking the integron ends. The genome annotations were analyzed by PATRIC 3.5.43 and confirmed by BLAST.

### Molecular Typing

For 33 clinical isolates of *S.* Typhimurium with class 1 integrons (named as IntI1-Col0 to IntI1-Col7), an MLST scheme with 7 housekeeping genes (*dnaN, sucA, purE, aroC, thrA, hemD, hisD*) was used to determine the sequence type (ST) (Larsen *et al*., 2012). Three isolates were evaluated by PCR and Sanger sequencing and the remaining 30 isolates were analyzed by WGS. The ST number was determined by comparing the allele sequence in the Center for Genomic Epidemiology database with MLST 2.0 (https://cge.cbs.dtu.dk/services/MLST/) and PubMLST (Public database for molecular typing and microbial genome diversity, https://pubmlst.org/bigsdb?db=pubmlst_salmonella_seqdef).

### Phylogeny by the SNP procedure

An analysis of phylogenies inferred from Single Nucleotide Polymorphism (SNPs) (Kaas *et al*., 2014) was performed at the Center for Genomic Epidemiology with CSI Phylogeny 1.4 (https://cge.cbs.dtu.dk/services/CSIPhylogeny/), using the WGS of 30 *S*. Typhimurium isolates positive for class 1 integrons (Table S1), and the reference genome *S*. Typhimurium LT2, AC AE006468.2 (McClelland *et al*., 2001). The WGS sequences were input in FASTQ format. The phylogenetic tree was performed under the recommended default parameters. The graphical viewer of phylogenetic tree was FigTree v1.4.4 2006-2018 (http://tree.bio.ed.ac.uk/software/figtree).

### Visualization and exploration of isolates containing class 1 integrons

The phylogenetic relationships between *S.* Typhimurium genomes with class 1 integrons as a tree of phylogenies inferred by SNPs is used as a framework to associate with relevant data such as class 1 integrons types, department (latitude, longitude), temporal (year of isolation), source, phenotypic resistance profile, Inc groups and MLST (https://microreact.org/project/vzZ3D_fds), we use Microreact (Argimón *et al*., 2016) is available free of charge (at http://microreact.org). The data file with the colors for the variables in comma separated value format (.CVS) and the Tree file in Newick format (.nwk) were input according to the Microreact instructions.

### Statistical Analysis

Binary logistic regression was used to investigate the effect of the source (independent variable) as a predictor of obtaining the class 1 integron genetic marker (dependent variable). The analysis included the three most frequent sources (blood culture, stool, swine carcass meat). The statistical model evaluated the null hypothesis “there is an association between the source and the presence of class 1 integron”. The critical value of 0.05 was used to predict whether to reject the null hypothesis. To look at the relationship between the dependent and independent variables, the Chi-square test of independence was performed, following the Hosmer and Lemeshow criterion. The weights of the variables were calculated by the Enter method. The Nagelkerke R-squared and the goodness-of-fit testthe Hosmer and Lemeshow test were used to determine the adequacy of the model. Statistical analysis was running in SPSS (Ferrán Aranaz, 1996).

## Results

### Colombian *S.* Typhimurium isolates positive for class 1 integrons

In this study, 39.03% (n = 153/392) of the clinical (stool and blood samples) and 22% (n=11/50) of the swine *S*. Typhimurium isolates harbored a complete class 1 integron. The presence of genes *qacE*Δ1 and *sul1* were confirmed by PCR amplification and DNA sequencing. Twelve different cassette arrays that share some family resistance genes were described, named IntI1-Col0 to IntI1-Col7 and IntI1-Col10 to IntI1-Col12. All isolates carried one type of class 1 integron, except for one isolate (GMR-S-125/2002) that contained two.

The most frequent cassette array from clinical samples was *dfr*7-*aac*-*bla*_OXA-2_ (named IntI1-Col1) found in 75.2% (n = 115/153). IntI1-Col1 was not identified in pork isolates. Other cassettes/cassette combinations only observed in clinical isolates were *aadA*2 (n = 9 isolates), *aadA1*2 (n = 1 isolate), *bla*_PSE-1_ (n = 4 isolates), *dfrA*17-*aadA*5 (n = 1 isolate), *dfrA*12-hypothetical protein-*aadA*2 (n = 1 isolate), and hypothetical protein-*aacA*38-Tryptophan synthase-*bla*_CTX-M-12_-mobile element protein-*bla*_OXA-1_ (n = 3 isolates), whereas the cassette arrays *aadA*7 (n = 2 isolates), *dfrA*12-*aadA*2 (n = 1 isolate) and *aac*-*bla*_OXA-2_ (n = 1 isolate) only occurred in pork isolates. The nomenclature for each class 1 integron (IntI1-Col0 to IntI1-Col12) and the cassette array described in this study are in Table 2 and Figure 1. In addition, an empty class 1 integron without variable region named IntI1-Col0 was found in 13.1% (n= 20/153) of the clinical isolates (Table S1, Table S3).

**Figure 1.**
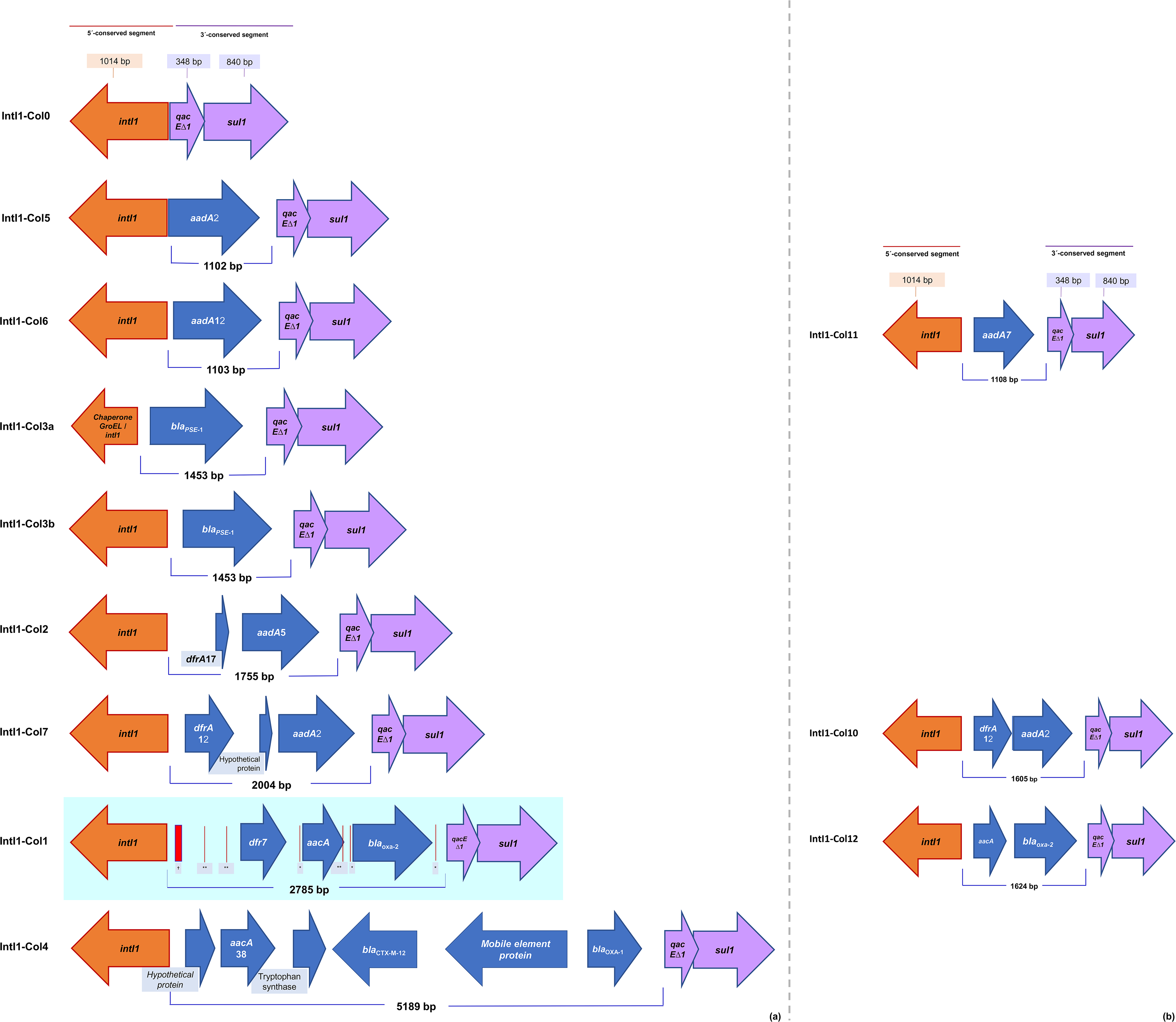
Schematic representation of class 1 integrons (IntI1-Col0 to Col12) with different cassette arrays of *S.* Typhimurium in Colombia (Col), 1997-2017. These integrons are aligned with respect to the class 1 integron integrase gene (*intI1*). The arrows indicating the orientation of transcription. The orange and purple arrows indicate the 5 ‘and 3’ conserved sequence respectively of the class 1 integrons groups, all gene cassettes in the variable region are presented as blue arrows. The predominant integron is seen in the blue box with the primary recombination site (attI1, thick red bar †), each cassette bounded by secondary recombination sites (slim red bar with the 1L-*attC* sequence, * and the 1R-*attC* sequence, **). (a) Class 1 integrons in clinical isolates, in (b) Class 1 integrons in swine isolates.

**Table 2.** Class 1 integrons in isolates of *S*. Typhimurium, Colombia 1997-2017. The Class 1 integrons of clinical samples were Col0 to Col7 (total positives = 153), and the class 1 integrons of swine samples were Col10 to Col12 (total positives = 11).

| Class 1 integron<sup>*</sup> | Variable region size (bp) | Variable region | n (%) |
| --- | --- | --- | --- |
| IntI1-Col<sup>Δ</sup>0 | No data | null variable region | 20 (13.1) |
| IntI1-Col1 | 2785 | *dfr7-aac-bla<sub>OXA-2</sub>* | 115 (75.2) |
| IntI1-Col2 / Col3a | 1755 / 1453 | *dfrA17-aadA5 / bla<sub>PSE-1</sub>* | 1 (0.65) |
| IntI1-Col3b | 1453 | *bla<sub>PSE-1</sub>* | 3 (2.0) |
| IntI1-Col4 | 5189 | Hypothetical protein<br>-*aacA38*-Tryptophan<br>synthase-*bla<sub>CTX-M-12</sub>*-Mobile<br>element protein-*bla<sub>OXA-1</sub>* | 3 (2.0) |
| IntI1-Col5 | 1102 | *aadA2* | 9 (5.9) |
| IntI1-Col6 | 1103 | *aadA12* | 1 (0.65) |
| IntI1-Col7 | 2004 | *dfrA12*-Hypothetical<br>protein-*aadA2* | 1 (0.65) |
| IntI1-Col10 | 1605 | *dfrA12-aadA2* | 1 (9.1) |
| IntI1-Col11 | 1108 | *aadA7* | 2 (18.2) |
| IntI1-Col12 | 1624 | *aac-bla<sub>OXA-2</sub>* | 1 (9.1) |

<sup>\*</sup>IntI1: Class 1 integron.
<sup>Δ</sup>Col: Colombia.
<sup>bp</sup>: base pairs.

Genes present in the variable region of the integrons such as *dfrA*, *aadA*, *aac*, *bla* confers resistance to several antimicrobial families such as trimethoprim-sulfamethoxazole (SXT), streptomycin (STR) and spectinomycin (SPT), ampicillin (AMP) carbenicillin (Cb), kanamycin (KAN), netilmicin (NET) tobramycin (TOB), cephalosporins (Cp), expanded spectrum cephalosporins as cefuroxime (CXM) and monobactam (M). Consequently, the isolates carrying an integron class 1 were associated with 14 different antimicrobial resistance profiles (with resistances to 1 to 5 antimicrobial families) (Figure S1). Almost half of the class 1 integron from clinical isolates were resistant to AMP-STR-SXT-TET (52.3%; n = 80/153). The second most frequent antimicrobial resistance profile was AMP-SXT-TET present in 17% (n = 26/153), followed by AMP-CHL-STR-SXT-TET (10.5%, n = 16/153) (Table S1).

In swine isolates, the class 1 integron were distributed within 3 antimicrobial resistance profiles (resistant to 1 to 3 families). The isolate carrying integron IntI1-Col10 exhibited AZT-SXT-TOB resistance profile, the isolates with IntI1-Col11 were resistant to CXM-TET and SXT, and the one with IntI1-Col12 also was resistant to CXM-TET (Table S1, Table S2).

Presence of class 1 integron was frequent in stool samples. In the 153 clinical isolates, class 1 integron were found in 112 stool samples (81.75%; n = 112/137) and 27 blood samples (11.4%, n = 27/237), and the remaining 14 were identified in other sources (100%, n = 4/4 of food and 71.43%, n = 10/14 of other samples). IntI1-Col2 and IntI1-Col3a were exclusively found in stool isolates, while IntI1-Col3b, IntI1-Col5, IntI1-Col6, and IntI1-Col7 were only found in blood isolates. IntI1-Col0, IntI1-Col1 and IntI1-Col4 were identified in 126 isolates obtained from stool, blood, and secretion samples (Figure 2).

**Figure 2.**
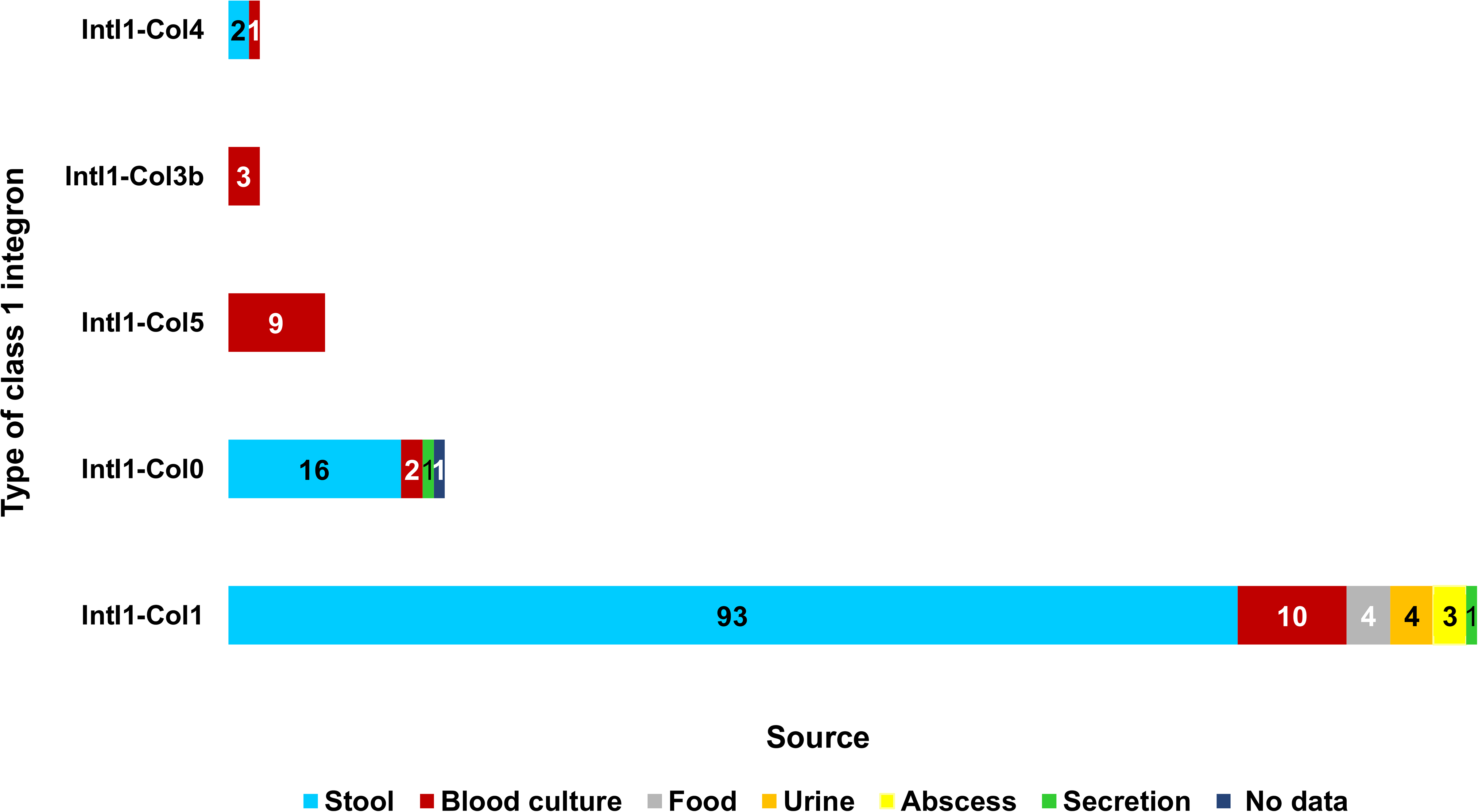
Class 1 integrons (n ≥ 3) and source in *S*. Typhimurium isolates from Colombian clinical samples, during 1997-2017. Stacked bar chart showing class 1 integrons types with frequencies ≥ 3 and the source from which the isolate was obtained. The X-axis shows the number of isolates of each integron type, the values within the bars represent the number of isolates for each source in each type of integron.

Among the class 1 integron found in swine isolates, 9.4% (n= 3/32) were recovered from the slaughterhouse isolates, followed by 12.5% (n= 1/8) obtained from the meat plant waste isolates. IntI1-Col10 was found in cutting plants isolates (25%, n= 1/4 of meat carcasses), IntI1-Col11 and IntI1-Col12 were found in slaughterhouse isolates (both integrons in 10.5%, n= 2/19 of meat carcasses; and another IntI1-Col11 isolate obtained from a knife in 25%, n= 1/4).

### IntI1-Col1 was the integron predominant in Colombian *S.* Typhimurium isolates

Despite the cassette array variability observed, one cassette array was predominant in clinical isolates included in this study. IntI1-Col1 was found in 75.2% (n= 115/153) of the isolates positives for class 1 integron. The unusual variable region flanked by the 5’-CS and 3’-CS is 2785 bp, start with a non-coding region of 689 bp containing a short segment of the *bla*_IMP-13_ gene (123 bp), followed by a *dfr*7, a partial *aac* gene and *bla*_OXA-2_ gene. Surprisingly, BLASTn analysis of the unusual 2785 bp displayed 100% sequence identity with five sequences reported in GenBank: 3 *Escherichia coli* isolates deposited in the CDC & FDA Antibiotic Resistance Isolate Bank (Accession Number AC: CP021722.1, CP021535.1 and CP020048.1) and with a *S*. Typhimurium (AC: AM237806.1) and *S*. Anatum (AC: AM237807.1) isolates recovered from food on the north coast of Colombia (O’Mahony *et al*., 2006).

A correlation was observed between the gene cassettes in the IntI1-Col1 and the resistance profile in the 115 clinical isolates. 96% isolates (n=110) were multidrug resistant with profile AMP-STR-SXT-TET observed in 60% (n= 69/115) of isolates, followed by AMP-SXT-TET in 20.9% (n= 24/115) isolates, and AMP-CHL-STR-SXT-TET in 8.7% (n= 10/115) isolates. Less common were AMK-STR and AMK identified in 3 and 2 isolates, respectively (Figure S1).

In addition, IntI1-Col1 appears to be broadly spread in the country. We found IntI1-Col1 in 12 departments (regions) from Nariño, located at the west of the country, to Bolivar in the north coast, and expand to Norte de Santander at the north bordering with Venezuela. IntI1-Col1 was mainly recovered in Antioquia (42.6%, n= 49/115), the Capital District of Bogotá (39.1%, n= 45/115) and Valle del Cauca (8.7%, n= 10/115). *S.* Typhimurium with IntI1-Col1 was identified as circulating in Colombia for 18 years, since 1997 until 2016, range of years selected for this study (Figure 3, Table S1).

**Figure 3.**
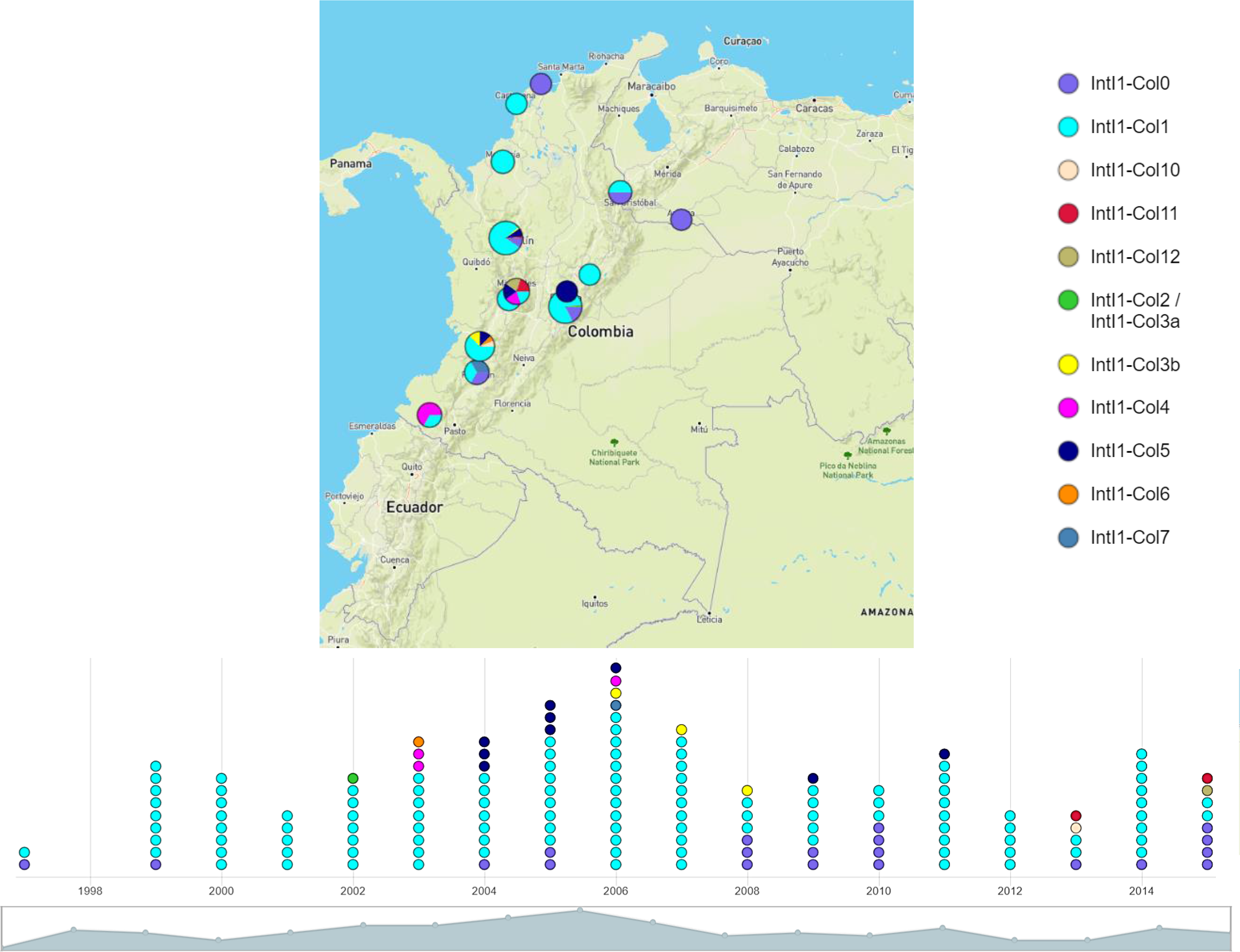
Spatial and temporal distribution of class 1 integrons in *S*. Typhimurium from Colombia (Col) between 1997-2017. The spatial and temporal visualization of class 1 integrons was elaborated on Microreact (DOI: 10.1099/mgen.0.000093). The top view panel shows the geographical distribution of class 1 integrons. The size of the circles matches the number of integrons identified in each department. In the lower view panel, the timeline shows the presence of all class 1 integrons since 1997, outstanding the predominance of IntI1-Col1 in aquamarine color. In the isolates analyzed from 1998, no class 1 integrons were identified.

### Several mobile elements are associated with Colombian IntI1

To understand the potential for lateral transfer of class 1 integron described in this study, we identified the main resistance plasmids and reconstructed the surrounding regions around the integrons by PCR and WGS. We identified seven common Enterobacterial incompatibility group (Inc) related with resistance determinants, including IncA/C, IncX1, IncHI1/HI2, IncI1 and ColpVC. The *S.* Typhimurium virulence plasmid pSTV with the IncF group was also identified in 34 out of 157 isolates carrying class 1 integron.

Interestingly, the predominant group was IncA/C (66 out of 104 isolates), followed by IncX1 (40 out of 104 isolates), IncF (34 out of 104 isolates), and small plasmids like ColpVC present in 11 out of 104 isolates. No relationship was observed between type of plasmids and type of class 1 integron, isolates with the predominant Int1-Col1 exhibited several plasmid combinations (Figure S2).

Isolates obtained from non-systemic sources (stool, urine, abscess, secretion) carried more plasmids of groups IncA/C, IncX1, IncF, IncI1, ColpVC, and IncHI1, than from systemic sources (blood) (Table 3). Plasmid groups IncA/C and IncA/C-IncX1 were identified in the isolates recovered from food and meat carcass of swine samples (Figure S3).

**Table 3.** Inc groups of plasmids and the source in S. Typhimurium of Colombia, 1997-2017. In stool samples, the IncA/C and IncX1 plasmids are more frequent and in blood culture samples the IncF plasmid is the predominant. Food isolates carry the plasmid groups IncA/C and IncA/C-IncX1.

| Fuente | IncA/C | IncF | IncX1 | IncA/C-ColpVC | IncA/C-IncF | IncA/C-IncX1 | IncF-IncI1 | IncF-IncX1 | IncA/C-ColpVC-IncX1 | IncF-IncHI2-IncX1 | IncA/C-IncF-IncI1 | IncA/C-IncF-IncX1 | IncA/C-ColpVC-IncHI1-IncX1 | IncA/C-ColpVC-IncF-IncX1 | IncA/C-ColpVC-IncF-IncHI1-IncI1 | IncA/C-ColpVC-IncF-IncHI1-IncI1-IncX1 |
| --- | --- | --- | --- | --- | --- | --- | --- | --- | --- | --- | --- | --- | --- | --- | --- | --- |
| Stool | 33 | 2 | 15 | 1 | 2 | 11 |  |  |  |  | 1 | 1 | 1 | 1 | 1 | 1 |
| Blood<br>culture |  | 16 |  | 2 | 3 |  | 1 | 1 | 1 |  |  |  |  | 2 |  |  |
| Food | 1 |  |  |  |  | 1 |  |  |  |  |  |  |  |  |  |  |
| Urine |  |  | 1 |  |  |  | 1 |  |  |  |  |  |  |  |  |  |
| Abscess | 1 |  | 1 |  |  |  |  |  |  |  |  |  |  |  |  |  |
| Meat<br>carcass<br>(swine) | 1 |  |  |  |  |  |  |  |  |  |  |  |  |  |  |  |
| No data |  |  |  |  |  |  |  |  |  | 1 |  |  |  |  |  |  |
| **Total** | **36** | **18** | **17** | **3** | **5** | **12** | **2** | **1** | **1** | **1** | **1** | **1** | **1** | **3** | **1** | **1** |

### WGS analysis of the genes surrounding class 1 integron showed two mechanisms of dispersion in clinical *S*. Typhimurium isolates

We confirmed that isolates with IntI1-Col2, IntI1-Col3a, IntI1-Col3b, and IntI1-Col5 belongs to the *Salmonella* genomic island 1 (SGI1) variant groups of the *S*. Typhimurium DT104 clone. IntI1-Col2 and IntI1-Col3a that are in the same isolate, where identified in separate contigs within the same isolate. A detail analysis of both contigs showed an SGI1 variant composed by the genes of In104 as *sgiT* toxin/antitoxin and SO15 to SO24, the presence of *floR*, *tetR* and *tetG* resistance genes and byIntI1-Col2 with an arrangement reported in *Proteus mirabilis* (Figure 1, Figure 4).

**Figure 4.**
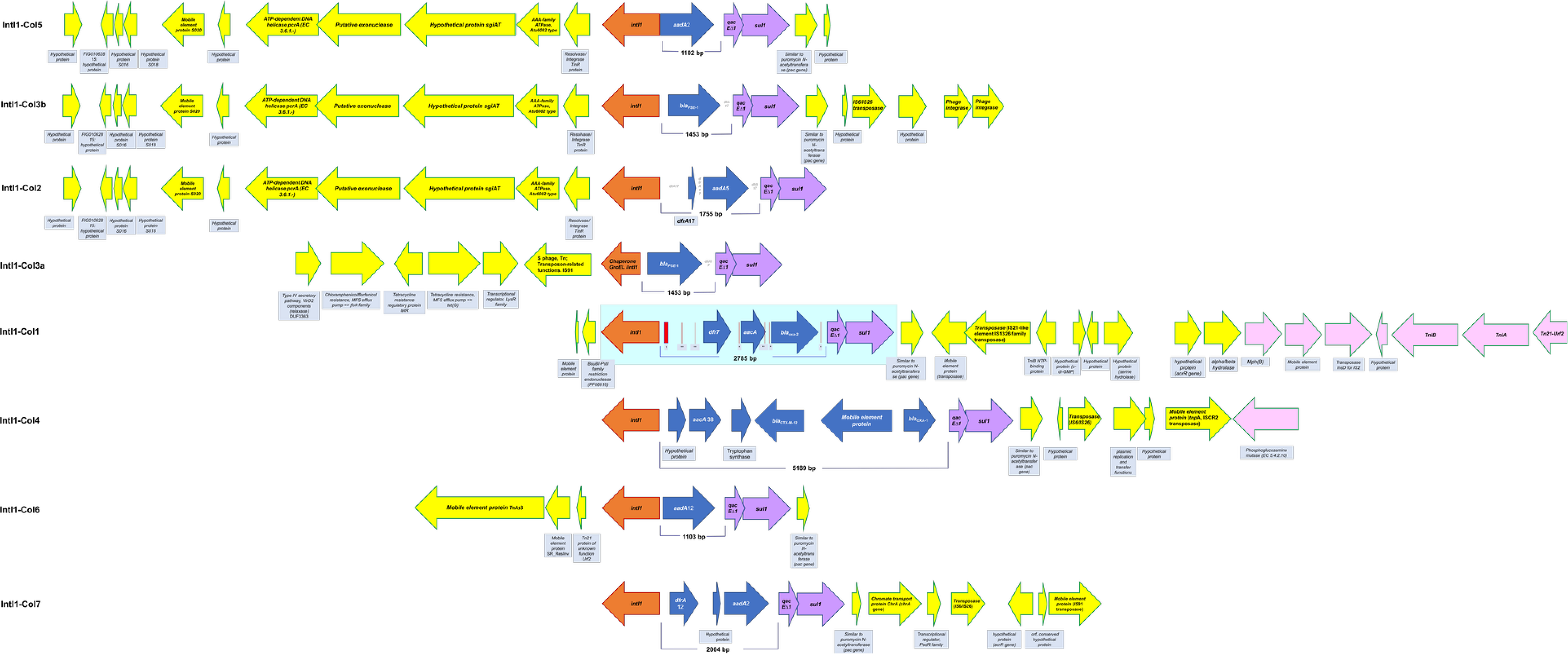
Schematic representation of surrounding regions in class 1 integrons (IntI1-Col1 to Col7) of Colombian *S.* Typhimurium, 1997-2017. The surrounding region of class 1 integrons are represented by yellow arrows, on similar mobile platforms, the synteny of the genes is evident. The integrons are represented by orange, blue and purple arrows and they are in line with the class 1 integron integrase gene (*intI1*). The arrows indicating the orientation of transcription.

IntI1-Col3b and IntI1-Col5 were identified in one contig per isolate by WGS. The presence of two characteristics class 1 integrons of the In104 element was confirmed by PCR of the variable region in which two bands of 1 and 1.5 kb were observed (in 3 isolates: IntI1-Col3b, n = 1 and IntI1-Col5, n = 2). These fragments were not sequenced.

The predominant IntI1-Col1 integron was inserted in a Tn5053 transposon flanked by Tn21 at the 3’-end, in one contig, as the reported previously (Kholodii *et al*., 1995, 1993). Genes located upstream of Intl1-Col1 were no identified due to the integrase gene at the start of the contig.

Gene context for IntI1-Col4, IntI1-Col6, and IntI1-Col7 was determined by association with other transposable elements, like TnAs3 from Tn3 family transposase, and the insertion sequences IS6/IS26 and IS91 (Figure 4, Table S4).

These results suggest that class 1 integron in Colombia are disseminated by mobile elements often associated with antibiotic resistance genes. We observed two mechanisms of dissemination, one by clonal spread of the integrons inserted in genomic islands, and second probably by horizontal transfer inserted in resistance plasmids as IncA/C o IncX1.

### The dispersion mechanisms of class 1 integrons suggest that in the Colombian population of *S.* Typhimurium there are phylogenetic subgroups

Genomic relatedness was analyzed for a subgroup of 30 clinical *S*. Typhimurium isolates carried class 1 integron. An unrooted phylogenetic SNP tree was generated (Figure 5).

**Figure 5.**
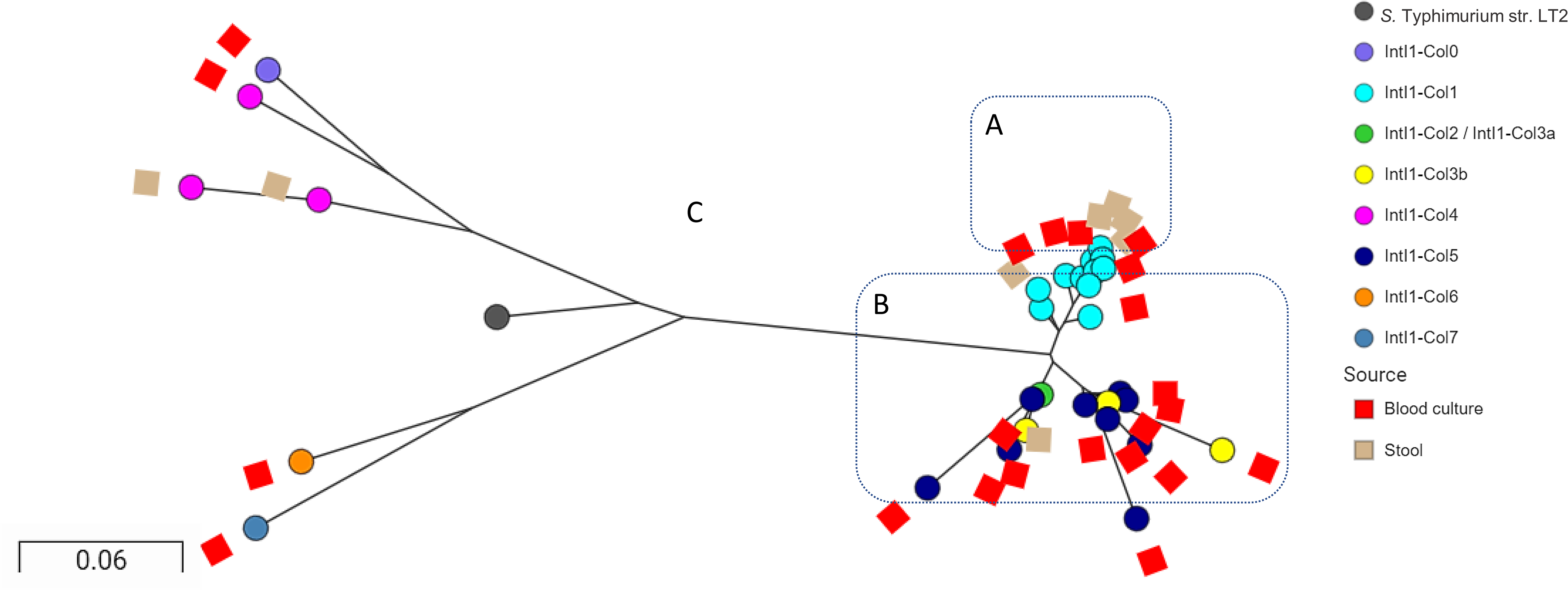
Phylogenetic tree by SNPs of 30 Colombian *S*. Typhimurium strains positive for class 1 integrons (IntI1), the separation of the clades allows linking the class 1 integrons types with the source. Cluster A consists of isolates with IntI1-Col1 from stool and blood sources, Cluster B groups the class 1 integrons mainly associated with blood culture. Cluster C with longer branches groups integrons that are mostly rare and present in blood and stool samples. The unrooted phylogenetic by SNPs was generated by the Center for Genomic Epidemiology with CSI Phylogeny 1.4 (https://cge.cbs.dtu.dk/services/CSIPhylogeny/) and visualized by Microreact (DOI: 10.1099/mgen.0.000093). The circles represent the types of integrons and the rectangles the source of the isolates. The length of each branch is proportional to the number of SNPs identified among the isolates.

Three phylogenetic clusters named A, B, and C were observed (Figure 5) in which clusters A and B were closely related, whereas cluster C was distant. Cluster A was composed exclusively of isolates with IntI1-Col1 inserted in Tn5053 obtained from stool and blood sources. Short branches in this group suggest a clonal relationship. Cluster B groups all the isolates with integrons present in SGI1 mainly associated with blood cultures, confirming the relationship between them. Cluster C grouped the isolates with class 1 integrons associated with other mobile elements recovered from stool and blood culture. However, the 30 clinical *S*. Typhimurium isolates were grouped by MLST into the ST19. These results confirm that the ST19 lineage is predominant in Colombian clinical *S.* Typhimurium isolates and suggests clonal subgroups according to the dispersal mechanism observed for class 1 integron.

## Discussion

In this work, we identified and characterized 12 different types of class 1 integron from clinical and swine resistant *S.* Typhimurium isolates in Colombia, most of them present in a low frequency. These result is agree with the abundant reports in the literature about the presence of several class 1 integron reported for this serovar (Antunes *et al*., 2006; Ranjbar *et al*., 2011; Vo *et al*., 2010). The class 1 integron described in this study maintain the complete structure (Stokes & Hall, 1989), and carry genes that encode resistance to common antimicrobial families (inhibitors of the folate pathway, aminoglycosides and β-lactams) (Firoozeh *et al*., 2012; Krauland *et al*., 2009). In the present study, there was a significant association between class 1 integron carriage and stool samples (odds ratio (OR) = 8.96), and among them the IntI1-Col2 and IntI1-Col3a are exclusive to this source. We also observed integrons only in blood culture samples as IntI1-Col3b, IntI1-Col5, IntI1-Col6 and IntI1-Col7 but with less frequency.

Despite the identification of 12 different types of class 1 integron, we found that IntI1-Col1 is the predominant in 75.2% of the isolates that carry on integrons and noteworthy this IntI1-Col1 is widely spread in the national territory for 18 years (1997-2017). The analysis of the 2.7 kb variable region of IntI1-Col1 showed an unusual array of genes, that start with a non-coding region of 689 bp, follow by a truncated aminoglycoside resistance gene and end with a functional beta-lactamase resistance gene (*bla*_OXA-2_), and isolates with this integron showed mainly the multidrug resistance profiles AMP-STR-SXT-TET, AMP-SXT-TET, and AMP-CHL-STR-SXT-TET.

In the non-coding region, we identified the complete attI1 region (65 bp) (Partridge *et al*., 2000), as well as two *attC* recombination sites that only conserve the 1R core site of *attC* instead the two conserved regions of *attC* (1L, 5’-GCCTAAC and 1R, GTTAGGC-3’) (Collis & Hall, 1992a; Recchia & Hall, 1995). The presence of two 1R core site of *attC* in the non-coding region could reflect excision events of two genes flanked at the 5’ end by the 1R core site (where the GTT triplet is located, a specific recombination site), one of which could be the *bla*_IMP-13_ gene, an excision reaction involving two *attC* elements (Collis *et al*., 1993). All these characteristics together could have affected the recombination capacity and therefore the ability of this integron to acquire, cleave, rearrange genes through site-specific recombination events described in these mobile genetic elements (Collis & Hall, 1992b; Collis *et al*., 1993) which could lead to this unusual variable region being conserved over time as part of the genetic platform of this class 1 integron.

BLAST analysis of the IntI1-Col1 sequence showed that it was reported in 2 *Salmonella* food isolates from Colombia (*S*. Typhimurium and *S*. Anatum) (O’Mahony *et al*., 2006). The presence of this integron in *Salmonella* isolated from food suggests circulation in different stages of the food chain. Integron IntI1-Col1 was also identified in 3 *E. coli* isolates from the USA (AC: CP021722.1, CP021535.1 and CP020048.1) and 1 *Salmonella enterica* [4,5,12: i: -] multidrug-resistant clinical isolate from Spain (Rodríguez *et al*., 2008), suggesting international circulation of IntI1-Col1 in *Salmonella* and other *Enterobacteriaceae*. However, IntI1-Col1 is believed to be endemic due to its prevalence in Colombian *S.* Typhimurium since 1997.

Knowing the class 1 integron present in Colombian *S.* Typhimurium isolates together with the resistance determinants they carry and recognize that among them there is a predominant integron (IntI1-Col1), provides tools to verify the effectiveness of the measures for the management and identification of cases of salmonellosis in the Colombian population, in travelers and other sources from the national territory.

We observed a 64 mg/mL ampicillin MIC value for two isolates with IntI1-Col1 (GMR-S-328/2007 and GMR-S-207/2008), that carry *bla*_OXA-2_ (data no show), a lower value than 1024 mg/mL reported by Antunes and Fisher (Antunes & Fisher, 2014). We hypothesized that the position of *bla*_OXA-2_ at the 3’-CS end of the variable region, was the indirect cause for the value MIC because are at the far end of the Pc promoter (last in the gene cassette arrangement) contrary to the common reports describe in *Salmonella* spp (Garrido *et al*., 2014; Montero *et al*., 2012) and in the INTEGRALL database, that show these gene in the 5’-CS initial region (Moura *et al*., 2009). However, this result is similar to report previously by Kadlec et al (Kadlec *et al*., 2007), who describe a class 1 integron only carry *bla*_OXA-2,_ suggesting that value MIC may be possible associated to be as part of integron.

In this study, we report for the first-time a less frequent integron (0.72%) identified as IntI1-Col4, with a variable region of 5189 bp, with genes that confer resistance to aminoglycosides and two different β-lactams genes (Figure 1). This integron was identified in the 3 Typhimurium clinical isolates that exhibited resistance to major number of antibiotic family (Table S1) and were recovered from stool and blood culture only in 2003 and 2006, after that year no more isolates with this characteristic were recovered in the surveillance.

Regarding the 10 others class 1 integron, the variable region revealed a predominance of genes that confer resistance to aminoglycosides (*aadA* and *aac* genes) alone or downstream of an trimethoprim resistance genes (*dfrA* genes), and upstream of β-lactams resistance genes (*bla*_OXA_, *bla*_CTX-M_ genes) and also the *bla*_PSE-1_ gene (Figure 1). The presence of these genes has been extensively documented as part of the class 1 integrons in *Salmonella* spp. (including *S*. Typhimurium) and Enterobacteriaceae, and usually correlated with the resistance phenotype observed in the respective isolates analyzed (Argüello *et al*., 2018; Krauland *et al*., 2009; Martinez-Freijo *et al*., 1998; Zhao *et al*., 2017). These characteristics make the class 1 integron identified as relevant elements in the acquisition, selection, maintenance, and dispersion of antibiotic resistance gene cassettes in the Colombian population of *S*. Typhimurium as previously noted for this serovar (Lopes *et al*., 2016).

An interesting result for us, describe in rare cases, is the presence of Int1-Col0 in 13.1% of the isolates. This is an empty integron without resistance genes in the variable region, that is an available platform for capture of resistance gene cassettes to generate new types of class 1 integrons in *S*. Typhimurium Colombian population, an adaptive advantage against selection pressure due to exposure to different antimicrobial agents (Firoozeh *et al*., 2019; Lapierre *et al*., 2008).

Regarding the integrons located in SGI-1 variants, the result are agree with that in Colombia circulated the pandemic *S.* Typhimurium MDR DT104 (Li *et al*., 2019), but with variations during the time. Surveillance of isolates associated with the *S*. Typhimurium MDR DT104 pandemic showed the same MDR signature of this phage type, which were also identified within the global time line in which it appeared and spread throughout the world (Leekitcharoenphon *et al*., 2016). We infer that in Colombia the first isolate derived from the Global epidemic *S*. Typhimurium MDR DT104 were replaced by other DT104 clones; thus the only isolate with IntI1-Col2 and IntI1-Col3 was collected in 2002, which was replaced by two DT104 clones, the first with IntI1-Col3b identified for a period of 3 years (2006-2008); and the second, a more successful clone carrying IntI1-Col5, was present for 8 years (2004-2011), which coincides with the period in which the Colombian cluster of *S.* Typhimurium MDR DT104 was identified (Li *et al*., 2019). The chromosomal location of these class 1 integrons means that there is a lower probability of horizontal transfer events of the antimicrobial resistance genes they carry (Zhang *et al*., 2018).

Regarding clonality, the 30 *S.* Typhimurium isolates belong to worldwide spread ST19 that circulates in Colombia (Li *et al*., 2019) and the SNP phylogenetic analysis suggests that the different mobile elements identified in this study affect the subpopulations lineages. We observed that the 11 *S*. Typhimurium isolates bearing IntI1-Col1 belong to a subclonal lineage (Figure 5A), whereas the isolates with SGI-1 grouped in another branch (Figure 5B). Our hypothesis is that mobile element Tn21-IntI1-Col1 is possibly located in the IncA/C plasmid, based on the result publish previously by ÓMahony (O’Mahony *et al*., 2006), and because literature mention that Tn21-Class 1 integron is a classical transposon insert in IncA/C plasmids in *S.* Typhimurium and other *Enterobacteriaceae* (Tamamura *et al*., 2013). This couple of elements are widespread among *Enterobacteriaceae* and they are recognize as elements of importance in public health, given their ability to confer multidrug resistance in human and animal pathogens (Fernández-Alarcón *et al*., 2011).

The above observations allows us to hypothesize that the dissemination of this Int1-Col1 in Colombia is a non-excluding two-way process, the first by horizontal transfer of the Tn21 in which the IntI1-Col1 is incorporated, possibly linked to the IncA/C plasmid, which allowed it to mobilize towards strains without these mobile genetic elements; and the second through vertical transfer that leads to the continuous dissemination of IntI1-Col1, and therefore to its establishment in the population (Domingues et al., 2012; Loftie-Eaton et al., 2016; Meng et al., 2011; Turner et al., 1998).

In the class 1 integrons identified in the Colombian swine industry samples, two were reported previously in similar sources in other countries. The IntI1-Col10 was described in *Salmonella* spp. (including *S*. Typhimurium) recovered from food products of animal origin (pig, chicken, duck) (Li et al., 2013), and in *E. coli* from clinical isolates (Shin *et al*., 2015). IntI1-Col11 is present in *E. coli* isolates from animals (Ajiboye et al., 2009; Mazel et al., 2000) and *Salmonella* spp. of broilers (Zhao et al., 2020).

Interestingly, we observed that IntI1-Col12 is like Int1-Col1 from clinical samples and was related with the presence of IncA/C plasmid in the same isolate. Int1-Col12 lack of all the 3’CS non-coding region and *dfr7* gene and carry the same truncated *aacA* and *bla*_OXA-2_ genes observed in IntI1-Col1. These similarities let us to consider that we are probably observed an integron evolution process in our population. We hypothesized two scenarios from the absence of the non-coding region and *dfr*7 gene. One, in swine industry isolates, could happen a selective pressure of antibiotics such as aminoglycosides and β-lactams, causing the rearrangement of the gene cassette mediated by the integrase *intI1* in where the *dfr*7 gene cassette would have been cleaved (Barraud & Ploy, 2015; Hocquet et al., 2012). Second, in clinical isolates, the insertion of the circularized *dfr*7 gene cassette by site-specific recombination catalyzed by the *intI1* integrase (*intI*1 gene) acquired from another clinical strain (Hall et al., 1999), forming the cassette gene arrangement characteristic of IntI1-Col1, an integron that could increase the adaptation of *S.* Typhimurium to human pressure.

The selective pressure of antibiotics in the swine production chain in Colombia is mainly due to the non-therapeutic use and abuse of antibiotics. There are reports of contamination with traces of mainly β-lactams and aminoglycosides in environmental and food sources (Agudelo-Londoño *et al*., 2012; Arenas & Moreno Melo, 2018). Faecal (human, animal) contamination from the ground and surface water sources have allowed the release of antibiotics, pathogenic microorganisms, other drugs that could explain the abundance of antibiotic resistance genes associated with class 1 integrons in aquatic environments (Agramont *et al*., 2020; Agudelo-Londoño *et al*., 2012).

The similarity between IntI1-Col1 and IntI1-Col12 in isolates of human and animal origin, respectively, allows highlighting the importance of the animal production in the appearance and dispersion of *S.* Typhimurium MDR strains carrying class 1 integron through the consumption of contaminated foods of animal origin. Multiresistance of animal and human sources, supporting this possible transmission route (Ajiboye *et al*., 2009; Zhao *et al*., 2003); however, the data obtained in this study do not allow us to rule out that the direction of the transfer of class 1 integron has been from humans to animal production species since close contact between these species promotes exposure to IntI1-bearing bacteria through water, food, handling or practices implemented in production processes, assumption supported by the transfer of class 1 integrons from humans to wild animals (McDougall *et al*., 2019). Regardless of the direction of class 1 integron transmission, the presence and maintenance of these integrons in *S*. Typhimurium MDR could be the result of an adaptive benefit of the bacterium in the face of selective pressure due to exposure to the antibiotics, stimulating their dispersion (Krauland *et al*., 2009), which is of interest because of the impact it can have on the production chain, health, food safety in humans and animals; hence the global importance of the One Health approach for the management of antibiotic resistance (Centers for Disease Control and Prevention (U.S.), 2019).

## Conclusions

In this study, we characterized 12 class 1 integron in *S*. Typhimurium clinical and swine industry isolates. In the clinical isolates a relationship between presence of class 1 integron and stool samples was observed. Among them, IntI1-Col1 inserted in Tn21 were the predominant in the country by 20 years and IntI1-Col4 was reported for the first time. *S*. Typhimurium isolates with these integrons were typed as ST19 mainly from non-systemic sources (stool) and with the IncA/C plasmid.

Two different insertion sites for class 1 integron were describe, one was chromosomal and the other in plasmids, as well as several transposons and insertion sequences surrounding the class 1 integron, exhibited several mechanisms of dispersion for these elements. The meaning of founding several class 1 integron, with differences in the genomic environment, is that Colombian *S*. Typhimurium isolates are exposed to the dynamic acquisition of mobile element present in other Enterobacteriaceae and that disseminated by horizontal or clonal transfer.

## Data Availability

All data produced in the present work are contained in the manuscript

## Data availability

The data of positive isolates for class 1 integron are summarized in S1 Table, year, origin, source, counties, phenotypic resistance profile, gene cassette array, type of class 1 integron, the sequences in FASTA format of the 139 isolates are available in ENA, and NCBI short Read Archive under the accession numbers listed in S1 table (BioProject Number PRJNA527650, SRA Study Number SRP188631), and the remain 13 FASTA sequences are available in at NCBI short Read Archive under the accession numbers listed in S1 table (BioProject Number SAMN25227165).

## Ethics approval and consent to participate

This research has the approval of the Technical Research Committee (CTIN) and the Ethics Research Committee (CEIN) of the Instituto Nacional de Salud (INS), Colombia with the codes CTIN-05-2015 and CTIN-05-2017. Clinical and patient ID from whom the *S*. Typhimurium clinical isolates were recovered, were anonymized.

## Competing interests

The authors declare that they have no competing interests.

## Authors’ contribution

N.Y.F-D. performed the laboratory tests, analysis of results, elaboration of tables, figures and drafted the manuscript. Y.L. performed the WGS experiments. E.N.U. performed the laboratory test. B.P-S. made the WGS experiments. E.L.O-R. performed lab tests. L.A.M. performed the phenotypic analysis of the isolates. J.C.D.H. participated in the design and coordination of the WGS experiments. A.K.C-C. and I.C.C-T. collected the pork samples and performed laboratory tests. S.Z-B. performed the statistical analysis. J.M.V. and M.W. conceived and coordinated the study, participated in the standardization and execution of tests, discussion of results and revision of the manuscript. All authors read and approved the final manuscript.

## Acknowledgements

This work was supported by COLCIENCIAS (SIGP 210471250745), COLCIENCIAS (Project no. 120358635758), the Ministerio de Agricultura y Desarrollo Rural (MADR) Convenio no. 20150360, PUJ PP-ID: 00006737, PY-ID: 00006865 and by the internal funds of the National Institute of Health, Colombia. N.Y.F-D. is supported by the special agreement of scientific and technological cooperation no. 014/14 of 2014 between Antonio Nariño University and the National Institute of Health, Colombia. E.N.U.is supported by ICETEX Scholarship Reciprocity Program for Foreigners in Colombia. To Andres Montilla for his support with the WGS test of a sample. Genome sequencing was performed at the Earlham Institute as part of the 10KSG consortium which is supported by the Global Challenges Research Fund data and resources grant (BBS/OS/GC/000009D). Next-generation sequencing and library construction were delivered via the BBSRC National Capability in Genomics and Single Cell (BB/CCG1720/1) at Earlham Institute, by members of the Genomics Pipelines Group.

## Supplementary information

**Figure S1. Susceptibility profile and main types of class 1 integrons (n≥ 3) detected in *S*. Typhimurium isolates from Colombian clinical samples, during 1997-2017.** Stacked bar graph showing the resistance profiles (n =13) in which class 1 integrons types with frequencies ≥ 3 were observed. The X-axis shows the number of isolates for each resistance profile, the values inside the bars represent the number of isolates for each integron type.

**Figure S2. Class 1 integrons types and Inc groups of plasmids in *S.* Typhimurium from Colombia (Col), 1997-2017.** Stacked bar chart showing the class 1 integrons types and Inc groups of plasmids present in the isolates was obtained. The labels on each of the bars represent the number of isolates for each Inc groups of plasmids. The X-axis shows the number of isolates of each integron type.

**Figure S3. Inc groups of plasmids (n≥3) and source in *S*. Typhimurium isolates from Colombia, during 1997-2017.** Stacked bar chart showing Inc groups of plasmids with frequencies ≥ 3 and the source from which the isolate was obtained. The X-axis shows the number of isolates of each Inc groups of plasmids, the values within the bars represent the number of isolates for each source.

**Table S1. Colombian isolates of *S*. Typhimurium with class 1 integrons recovered between 1997 and 2017.** The table shows the characteristics of the *S*. Typhimurium isolates, the types of class 1 integrons they carry, and the gene array identified in their variable region.

**Table S2. Data of all swine samples of *S*. Typhimurium used.**

**Table S3. List of variable region genes of class 1 integrons in Colombian *S*. Typhimurium isolates with GenBank accession numbers and two Patric’s annotations for similar sequences.**

## Notes

### Competing Interest Statement

The authors have declared no competing interest.

### Author Declarations

This research has the approval of the Technical Research Committee (CTIN) and the Ethics Research Committee (CEIN) of the Instituto Nacional de Salud (INS), Colombia with the codes CTIN-05-2015 and CTIN-05-2017. Clinical and patient ID from whom the S. Typhimurium clinical isolates were recovered, were anonymized.

