## Supplementary material for "Class 1 integrons in clinical and swine industry isolates of *Salmonella* Typhimurium from Colombia, dating 1997 to 2017": FigureS1-S3

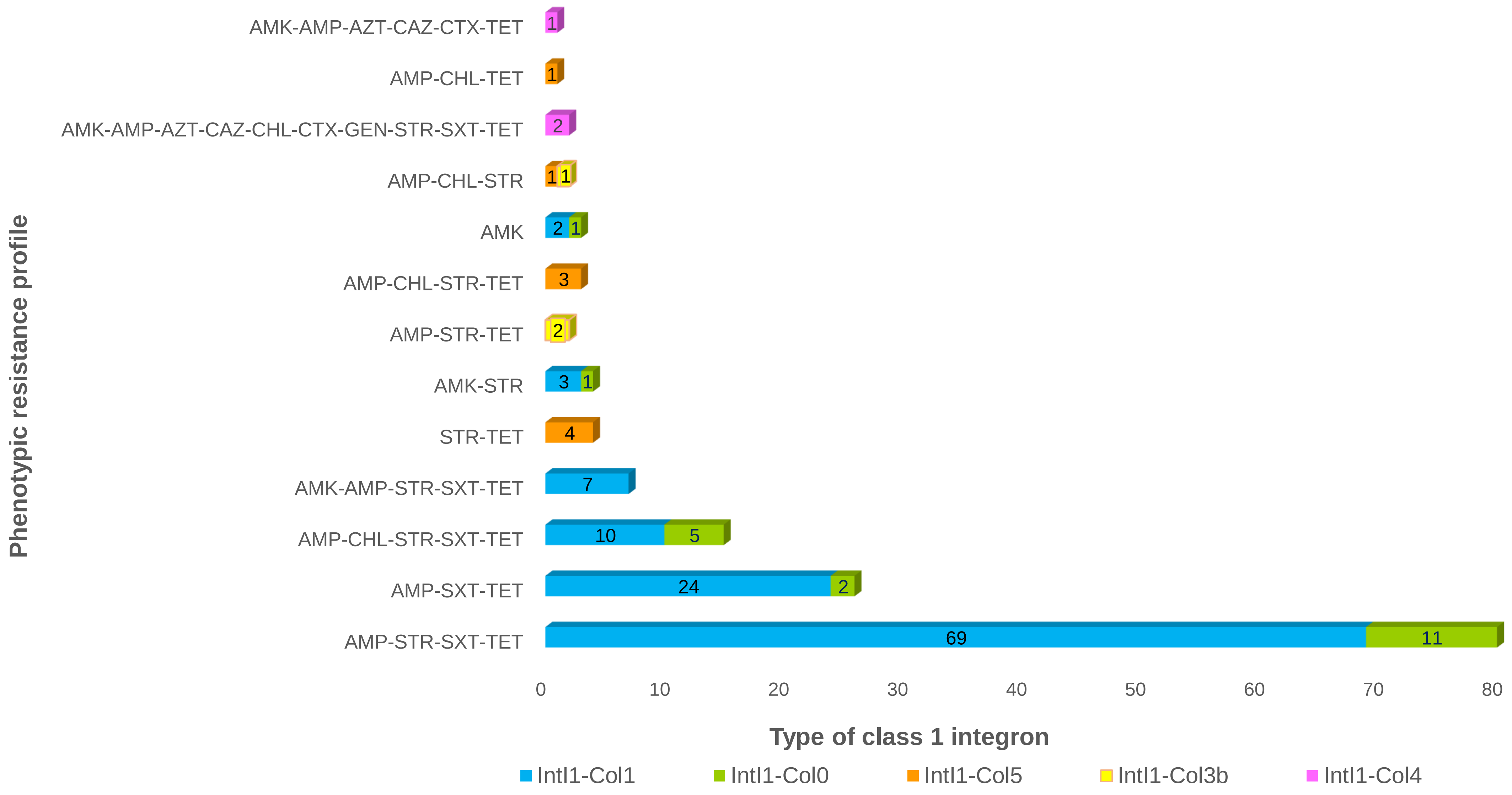


**Figure S1**. Susceptibility profile and main types of class 1 integrons (n≥ 3) detected in *S*. Typhimurium isolates from Colombian clinical samples, during 1997-2017.

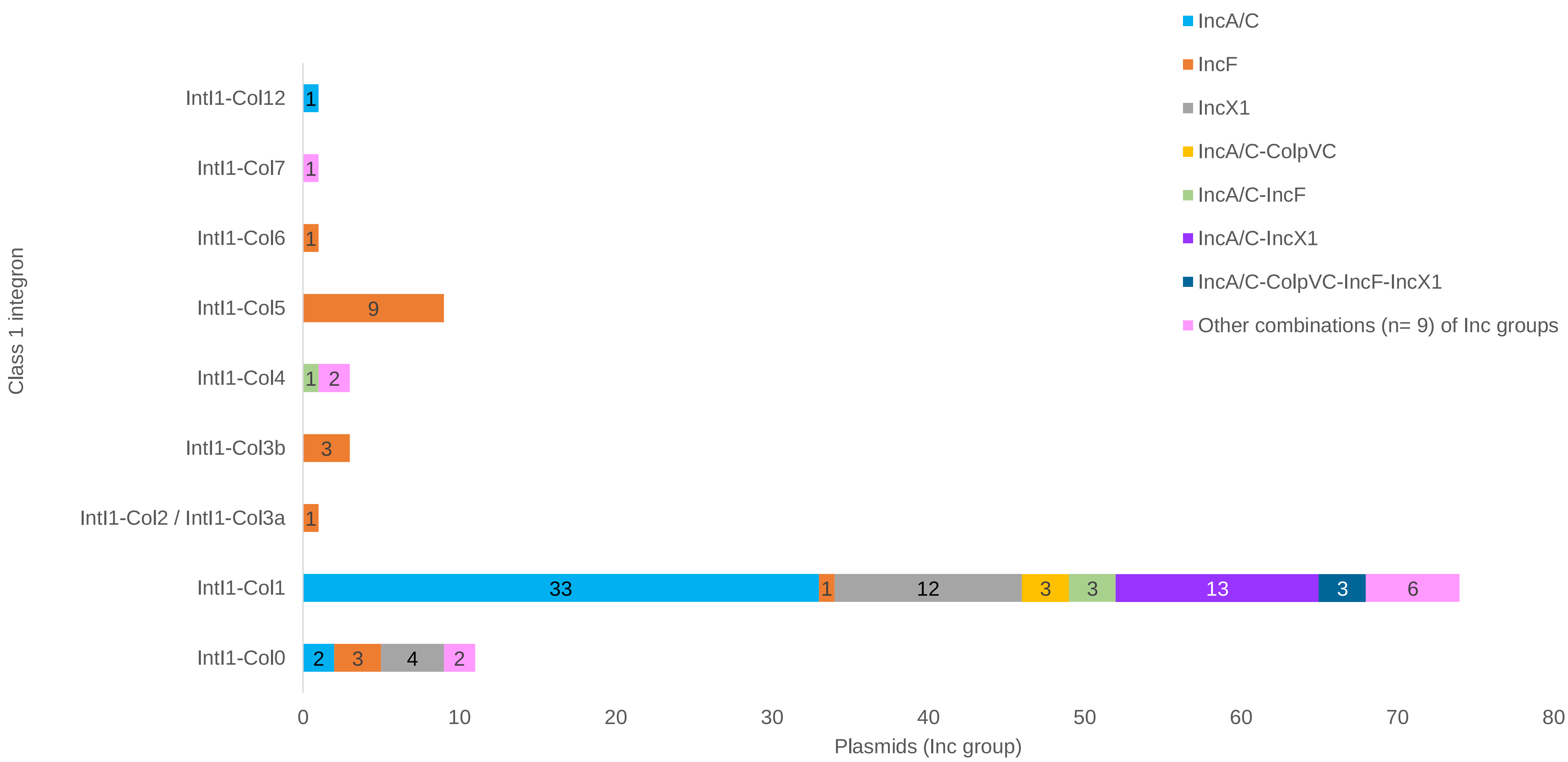


**Figure S2.** Class 1 integrons types and Inc groups of plasmids in *S*. Typhimurium from Colombia (Col), 1997-2017.

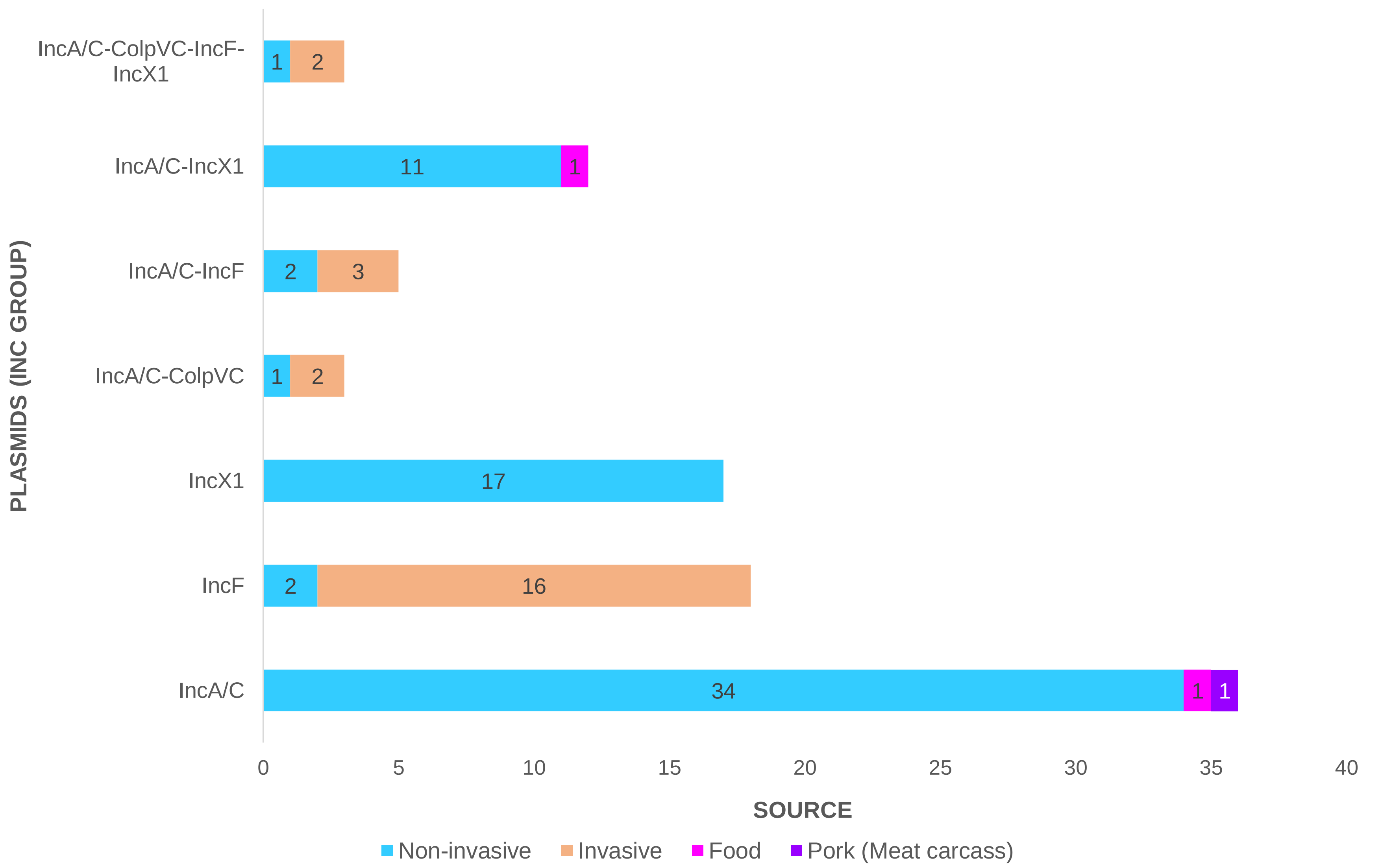


**Figure S3.** Inc groups of plasmids (n≥3) and source in *S.* Typhimurium isolates from Colombia, during 1997-2017.
